# Sociodemographic predictors of asthma control and exacerbation among children in Central Kenya: A hospital-based cross-sectional study

**DOI:** 10.64898/2026.02.05.26345704

**Authors:** Robai Linet, Mukala Joseph, Kanoi Bernard, Peter Mwaura, Ogweno Kennedy, Kamau David

**Affiliations:** Mount Kenya University, School of clinical medicine, Thika, Kenya; Mount Kenya University, Institute of tropical medicine, Thika, Kenya; Mount Kenya University, School of medicine, Thika, Kenya

**Author notes:** Corresponding author: Robai Linet.

**Keywords:** Asthma exacerbation, sociodemographic factors, caregiver education, children, Thika Level 5 Hospital, Kenya

## Abstract

**Introduction:** Childhood asthma remains a major public health challenge in low- and middle-income countries, where social and economic factors influence disease outcomes. This study examined the sociodemographic determinants of asthma exacerbation among children attending clinic at Thika Level 5 Hospital, Kenya.

**Methods:** This study employed a hospital-based cross-sectional study involving 108 caregivers– child dyads, each consisting of a child with confirmed asthma and their primary caregiver. The dyads were recruited from the pediatric asthma clinic from 31^st^ March 2025 to 30^th^ April 2025 then follow up was conducted for six months’ form 1^st^ May to 31st October 2025. Data were collected using structured questionnaires and clinic records. Descriptive statistics summarized sociodemographic characteristics, while chi-square tests and logistic regression assessed associations between caregiver factors and asthma control.

**Results:** The mean age of children was 8.1 years (range 3–17), with males comprising 57.4%. Most caregivers were mothers (88%), had secondary education (57.4%), and were in informal employment (75.9%). Household income was low for 59.3% of participants (<KES 30,000/month). Caregiver education (AOR=2.8; 95% CI:1.5–5.2; p=0.001) was the strongest predictor of asthma control, followed by medical insurance (χ^2^=10.41; p=0.001). Formal employment and higher income were significantly associated with controlled asthma (χ^2^=6.45; p=0.04 and χ^2^=9.72; p=0.02 respectively). Urban residence modified the positive effect of education on asthma management (interaction AOR=1.9; p=0.03).

**Conclusion:** Caregiver education level, employment, income, and medical insurance significantly influence asthma control among children. Enhancing health literacy and expanding insurance coverage under the Social Health Authority (SHA) can improve asthma outcomes in Kenyan children.

## Background information

Asthma is a chronic inflammatory disorder of the airways characterized by variable airflow obstruction and airway hyper responsiveness, affecting an estimated 262 million people globally and contributing to nearly half a million deaths annually (2). Despite being preventable and manageable, asthma remains a leading cause of disability-adjusted life years among children, significantly affecting school attendance, academic performance, and overall quality of life (3,4). Pediatric asthma accounts for a substantial burden on healthcare systems, with frequent hospitalizations and emergency department visits linked to poor control and environmental triggers (5,6).

In sub-Saharan Africa, the burden of asthma is increasing rapidly due to population growth, industrialization, environmental pollution, and inadequate access to essential asthma medications (7,8). The World Health Organization and Global Asthma Network have reported a growing prevalence in low- and middle-income countries where healthcare infrastructure and public awareness remain limited (4,9). Urbanization in Kenya and other African countries has introduced new risk exposures, including particulate matter from traffic, industrial emissions, and domestic biomass fuel smoke, which collectively exacerbate respiratory symptoms in vulnerable populations (16,17). Moreover, household environmental factors such as overcrowding, presence of pets, and poor ventilation further compound asthma morbidity (18). Recent surveys in Kenya estimate asthma prevalence among school-aged children to range from 10% to 14%, underscoring an urgent public health concern (6,25).

Socioeconomic and demographic determinants play pivotal roles in asthma management and clinical outcomes. Caregiver education, occupation, income level, and health insurance coverage influence access to healthcare services, medication adherence, and the ability to recognize early symptoms of exacerbation (14,15). Studies in Ethiopia, Ghana, and South Africa have consistently shown that caregivers with higher educational attainment are more likely to adhere to medication and proper inhaler use technique, implement trigger-avoidance measures, and seek timely medical interventions (10–12). Conversely, low socioeconomic status and limited health literacy contribute to underdiagnoses, delayed treatment, and frequent emergency visits (13,20,28).

In Kenya, initiatives such as the BReATHE (Beating Regional Asthma through Health Education) intervention, implemented at Thika Level 5 Hospital, have sought to strengthen caregiver awareness, correct inhaler use technique, and follow-up compliance among asthmatic children (8,24). However, there remains limited evidence quantifying how caregiver sociodemographic characteristics influence asthma control and exacerbation frequency in clinical settings. Understanding these determinants is critical for developing targeted interventions to reduce pediatric asthma morbidity and enhance outcomes under Kenya’s Universal Health Coverage (UHC) framework (21).

This study therefore analyzed the sociodemographic factors influencing asthma exacerbation among children attending Thika Level 5 Hospital, Kiambu County, Kenya, focusing on children’s age, gender, medical insurance cover and area of residence. Also caregiver’s occupation, education level and income.

## Methods

### Study design and setting

A hospital-based cross-sectional study was conducted between conducted from 31^st^ March, 2025 to 31^st^ October at Thika Level 5 Hospital, a major referral facility in Kiambu County serving both urban and peri-urban populations. Thika Level 5 Hospital is a county referral facility located in Thika Town, Kiambu County, approximately 40 kilometers northeast of Nairobi, along the Nairobi–Garissa highway. Administratively, it’s under Kiambu County Government and is one of the three Level 5 hospitals in the county (Ministry of Health, 2014). The hospital serves a catchment population of about 1.5 million people and provides specialized referral services to patients from Thika Sub-County and neighboring regions such as Murang’a, Machakos, Garissa, and Nairobi (Kiambu County Health Department, 2018). It is the busiest Level 5 hospital in Kiambu County, with a high patient load across both outpatient and inpatient departments (Thika Level 5 Hospital Annual Report, 2017). The study was conducted within the pediatrics department, which caters to both outpatient and inpatient pediatric populations. The pediatric outpatient clinic manages follow-up cases of children with chronic conditions such as asthma, while the inpatient pediatric wards admit children with acute conditions, including severe asthma exacerbations (Ministry of Health, 2016). In addition, Thika Town is a highly industrialized area where children are exposed to environmental risk factors such as air pollution and allergens, which have been shown to contribute to a higher prevalence and exacerbation of asthma (Mwaura et al., 2019; Gakidou et al., 2020). These characteristics make Thika Level 5 Hospital a suitable site for assessing the effectiveness of the BReATHE intervention in mitigating asthma exacerbation.

### Study population

Participants were 108 children aged 3–17 years known asthmatic and their primary caregivers. Eligibility included at least six months of follow-up in the hospital’s asthma clinic.

### Sampling procedures and techniques

A sample of 108 respondents was drawn from the total study population. According to the recent report (BReATHE Study Report, 2023), it was suggested that 7.5% of asthmatic children have no knowledge about asthma. For patients with knowledge, it was expected that there would be a 40% difference in the population with reduced asthmatic attacks. By using the two-sample means comparison formula, the required sample size was calculated

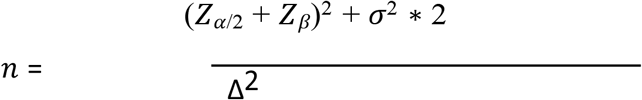

Where:

n is the sample size

*Z*_*α*/2_ is Z-value for the desired significance level (e.g1.96 for 5% significance level)

*Z*_*β*_ is Z-value for the desired power (e.g 0.84 for 80% power)

*σ*^2^ is population variance (or estimated variance from prior studies)

Δ^2^ is the minimum detectable difference between group means (Simba,2021).

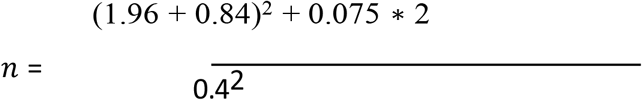

*n* = 49.9=49=49×2=98. 10% attrition rate=108

A minimum of 108 participants was targeted to include the attrition rate of 10%.

Although this manuscript focuses on predictors of asthma outcomes, the sample size was determined based on the primary outcome of the parent intervention study.

### Sampling technique

The study employed a stratified sampling approach, with age serving as the basis for stratification to ensure representation across different caregiver age groups. Within each stratum, participants were then recruited through simple random sampling, allowing every eligible caregiver an equal chance of being selected. The respondents who completed the questionnaire were caregivers of the affected children.

### Data collection tools

Semi structured questionnaires were developed to capture details on demographics, health patterns, and behaviors exhibited by caregivers and their children. In addition, standardized tools were incorporated to enhance reliability and comparability of the data. The Asthma Control Test (ACT) for children was used to assess asthma symptoms, control, and impact on daily activities, while the WHOQOL-BREF instrument was applied to measure caregiver quality of life across the physical, psychological, social, and environmental domains. These instruments were chosen because they are validated, widely used in similar studies, and suitable for both clinical and community settings.

### Data analysis

Data were entered and cleaned in SPSS version 31. Descriptive analysis summarized participants’ characteristics. Chi-square tests evaluated bivariate relationships, and binary logistic regression determined independent predictors of asthma control. Adjusted odds ratios (AOR) and 95% confidence intervals (CI) were computed, p < 0.05 was considered statistically significant.

### Ethical considerations

This study adhered to established ethical standards for research involving human participants. Ethical clearance was obtained from the Mount Kenya University Ethics Review Committee (ERC), after which a letter of introduction was issued to the National Commission for Science, Technology and Innovation (NACOSTI). A research permit was subsequently granted by NACOSTI (License No. NACOSTI/P/25/416394). The permit was presented to the Kiambu County Department of Health Research Unit, the County Director of Education, and the **County** Commissioner’s Office for authorization. Permission to conduct the study was then obtained from the Kiambu County Government and the Medical Superintendent of Thika Level 5 Hospital. Participant recruitment was conducted from 31^st^ March 2025 to 30^th^ April 2025 then follow up was done for six months’ form 1^st^ May to 31st October 2025. Prior to enrolment, parents or legal guardians of all participating minors were fully informed about the purpose of the study, the procedures involved, the voluntary nature of participation, the right to withdraw at any time without penalty, and the measures taken to protect participants’ privacy, anonymity, and confidentiality. Written informed consent was obtained from parents or legal guardians of all participating minors. Where applicable, assent was also obtained from children in accordance with ethical guidelines approved by the Ethics Review Committee.Participants were informed that the study did not provide any form of financial or material compensation, including transport reimbursement, and this information was clearly communicated during the consent process. To ensure confidentiality, participants were assigned unique identification codes rather than personal identifiers. Hard-copy data were securely stored in locked cabinets, while electronic data were password-protected and accessible only to the principal investigator.

## Results

**Table 1.** Sociodemographic characteristics of the participants (n=108)

| **Variable** | **Category** | **Frequency(n)** | **Percentage (%)** |
| --- | --- | --- | --- |
| **Child gender** | **Male** | **62** | **57.4** |
|  | **Female** | **46** | **42.6** |
| **Age group(years)** | **3-7** | **49** | **45.4** |
|  | **8-12** | **36** | **33.3** |
|  | **13-17** | **23** | **21.3** |
| **Caregiver Relation** | Mother | 95 | 88.0 |
|  | Father | 7 | 6.5 |
|  | Guardian | 6 | 5.5 |
| **Caregiver education** | Primary | 25 | 23.1 |
|  | Secondary | 62 | 57.4 |
|  | Tertiary | 21 | 19.5 |
| **Employment** | Formal | 26 | 24.1 |
|  | Informal | 87 | 75.9 |
| **Monthly income (KES)** | <KES 10,000 | 28 | 25.9 |
|  | 10,000–30,000 | 36 | 33.4 |
|  | >30,000 | 44 | 40.7 |
| **Residence** | Urban | 68 | 63.0 |
|  | Rural | 40 | 37.0 |
| **Medical cover** | Yes | 42 | 38.9 |
|  | No | 66 | 61.1 |

**Table 2.**
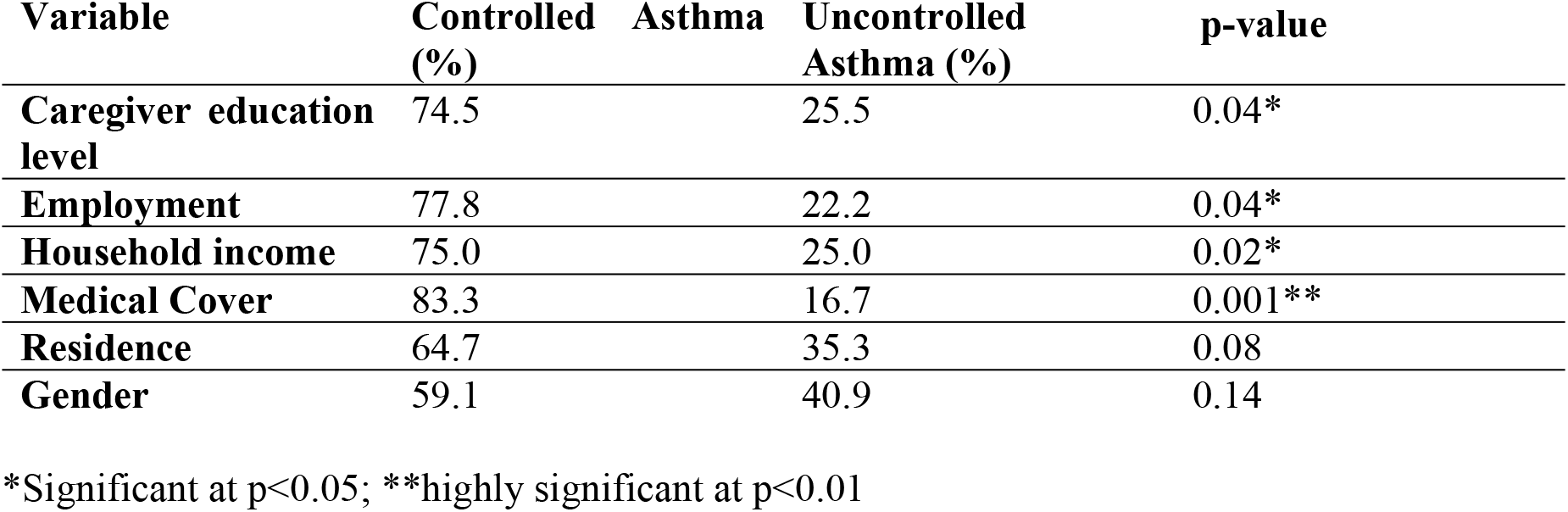
Association between sociodemographic factors and asthma control.

**Table 3.** Multivariate logistic regression predicting controlled asthma.

| Predictor | AOR | 95% CI | p-value |
| --- | --- | --- | --- |
| Caregiver Education (secondary or higher) | 2.8 | 1.5–5.2 | 0.001** |
| Formal Employment | 1.9 | 1.1–3.8 | 0.02* |
| Monthly Income > KES 30,000 | 2.3 | 1.2–4.1 | 0.01* |
| Medical Cover | 3.2 | 1.6–6.3 | 0.001** |
| Urban Residence × Education (interaction) | 1.9 | 1.1–3.2 | 0.03* |

\*Significant at $p < 0.05$ ; \*\*highly significant at $p < 0.01$

The interaction between urban residence and caregiver education was statistically significant (AOR = 1.9, 95% CI: 1.1–3.2, p = 0.03), indicating that the effect of higher caregiver education on asthma outcomes differs by place of residence. Interestingly, the protective effect of higher caregiver education was more pronounced in urban settings, suggesting that health literacy is crucial for navigating urban environmental triggers such as pollution, crowded housing, and limited green spaces. This implies that educational interventions may have an especially strong impact in urban areas.

## Discussion

This study demonstrates that sociodemographic factors significantly influence asthma control among children in central Kenya. The mean age of participants was 8.1 years, with a predominance of male children, reflecting global patterns where prepubertal male children have a higher prevalence of asthma compared to female children (3,5). Similar trends have been documented in other African countries, including Nigeria and Tanzania, suggesting a consistent gender predisposition across diverse populations (7,9,26).

Caregiver education level and occupation emerged as strong determinants of asthma control. Caregivers with higher education levels demonstrated better understanding of asthma medications, inhaler use techniques, trigger avoidance, and adherence to prescribed medication regimens, leading to improved asthma outcomes among their children. These findings are consistent with studies from Ethiopia and South Africa that identified caregiver literacy as a major predictor of asthma management success (10,11). Moreover, employment status was positively associated with asthma control, likely reflecting the influence of socioeconomic stability that enables caregivers to afford medications, transportation to clinics, and follow-up visits (8,15).

This relationship underscores the broader impact of economic empowerment on chronic disease management in low- and middle-income settings (14,29).Low income and lack of health insurance were also identified as key barriers to achieving optimal asthma control. Financial hardship restricts access to controller medications and routine healthcare visits, a finding consistent with studies in other low-resource contexts (20,21). In this study, only 38.9% of caregivers reported having medical insurance, highlighting a significant gap in financial protection for families managing chronic pediatric illnesses. Expansion of the Social Health Authority (SHA) to comprehensively cover pediatric asthma could help address these inequities and ensure sustained access to essential asthma care services (21,24).

The role of residential environment was more nuanced. Urban residence appeared to have both positive and negative effects on asthma outcomes. On one hand, it improved access to healthcare facilities and specialized asthma clinics; on the other, it increased exposure to air pollutants, dust, and allergens that exacerbate asthma symptoms (16,17,23). The observed positive interaction between urban residence and caregiver education level suggests that educated caregivers in urban areas may be better equipped to identify and mitigate environmental triggers as they have better understanding of asthma disease, thereby reducing exacerbations. This finding aligns with evidence from Ghana and Brazil, where higher education level was associated with increased asthma health literacy and among urban caregivers it correlated with better asthma control despite environmental challenges (12,13).

The implications of these findings are multifaceted. Effective asthma control among children requires not only biomedical interventions but also strategies that address sociodemographic determinants of health, such as caregiver education, income level, and living conditions. These factors influence treatment adherence, health-seeking behavior, and overall disease management (8,24). Strengthening community-based health education and improving access to care could substantially reduce the burden of pediatric asthma hospitalizations and exacerbations in Kenya. Overall, this study reinforces that caregiver education level, employment, income, and access to medical insurance are pivotal in determining asthma control among Kenyan children (8, 14, 20). Addressing these factors through multisector interventions including strengthening health literacy, promoting economic stability, and expanding universal health coverage could improve asthma management and quality of life for affected children. These results advocate for policy reforms to include pediatric asthma care within social health authority (SHA) benefits and underscore the importance of incorporating structured asthma education into both school health and community outreach programs (21, 24, 30).

## Data Availability

Data cannot be shared publicly because of confidentiality and data protection Act. Data are available from the Kennedy Odhiambo through the or researchers who meet the criteria for access to confidential data.

## Recommendations

### 1. Policy

Strengthen SHA to cover pediatric asthma follow-ups and medication refills and also to subsidies asthma medications.

### 2. Environmental Control

Reduce air pollution, especially in urban areas, to minimize asthma triggers and improve respiratory health among children.

### 3. Research

Conduct longitudinal studies exploring environmental and genetic modifiers of asthma control in Kenyan settings.

## Acknowledgements

The authors express their sincere gratitude to the management and staff of Thika Level 5 Hospital for their support during data collection. Special thanks go to the caregivers and children who participated in this study. The authors also acknowledge the Mount Kenya University, School of clinical medicine, for academic and ethical support. No external funding was received for this study, and the funders had no role in the design, data collection, analysis, interpretation, or writing of the manuscript.

## Authors’ contributions

RL is the principal investigator and overall owner of the study who conceived and designed it and wrote the manuscript. KO is research assistant and helped with data collection and analysis of data. JM, BK, PM and DK provided mentorship, guided data interpretation, and critically reviewed the manuscript for important intellectual content. All authors read and approved the final version of the manuscript.

